# Incidence of SARS-CoV-2 infection according to baseline antibody status in staff and residents of 100 Long Term Care Facilities (VIVALDI study)

**DOI:** 10.1101/2021.03.08.21253110

**Authors:** Maria Krutikov, Tom Palmer, Gokhan Tut, Chris Fuller, Madhumita Shrotri, Haydn Williams, Daniel Davies, Aidan Irwin-Singer, James Robson, Andrew Hayward, Paul Moss, Andrew Copas, Laura Shallcross

**Affiliations:** UCL Institute of Health Informatics, London, UK; UCL Institute for Global Health, London, UK; Institute of Immunology and Immunotherapy, University of Birmingham, Birmingham, UK; Public Health England; Four Seasons Healthcare Group, Wilmslow, Cheshire; Palantir Technologies UK Ltd, London, UK; Department of Health and Social Care, London, UK; UCL Institute of Epidemiology & Healthcare, London, UK; Health Data Research UK

**Author notes:** Correspondence to: Dr Laura Shallcross, UCL Institute of Health Informatics, 222 Euston Road, London NW1 2DA, UK., 0203 549 5540. These authors contributed equally to this work (Maria Krutikov, Tom Palmer, Gokhan Tut).

## Abstract

**Background:** SARS-CoV-2 infection represents a major challenge for Long Term Care Facilities (LTCFs) and many residents and staff are now sero-positive following persistent outbreaks. We investigated the relationship between the presence of SARS-CoV-2 specific antibodies and subsequent infection in this population.

**Methods:** Prospective cohort study of infection in staff and residents in 100 LTCFs in England between October 2020 and February 2021. Blood samples were collected at baseline (June 2020), 2 and 4 months and tested for IgG antibodies to nucleocapsid and spike protein. PCR testing for SARS-CoV-2 was undertaken weekly in staff and monthly in residents. The primary analysis estimated the relative hazard of a PCR-positive test by baseline antibody status, from Cox regression adjusted for age and gender, and stratified by LTCF.

**Findings:** Study inclusion criteria were met by 682 residents and 1429 staff. Baseline IgG antibodies to nucleocapsid were detected in 226 residents (33%) and 408 staff (29%). A total of 93 antibody-negative residents had a PCR-positive test (0.054 per month at risk) compared to 4 antibody-positive residents (0.007 per month at risk). There were 111 PCR-positive tests in antibody-negative staff (0.042 per month at risk) compared to 10 in antibody-positive staff (0.009 per month at risk). The adjusted hazard ratios for reinfection in staff and residents with a baseline positive versus negative antibody test were 0.13 (95% CI 0.05-0.40) and 0.39 ((95% CI: 0.19-0.77) respectively. Of 12 reinfected participants with data on symptoms, 11 were symptomatic. Antibody titres to spike and nucleocapsid were comparable in PCR-positive and PCR-negative cases.

**Interpretation:** The presence of IgG antibodies to nucleocapsid was associated with substantially reduced risk of reinfection in staff and residents for up to 10 months after primary infection.

**Funding:** UK Government Department of Health and Social Care

**Research in context:** *Evidence before this study:* We performed a systematic search of MEDLINE (Ovid) and MedRxiv on 18 January 2021 for studies in LTCFs that described the risk of infection in individuals who were seropositive for SARS-CoV-2 compared to individuals who were seronegative. Search terms were deliberately broad to improve capture of relevant literature and included “SARS-CoV-2”OR “COVID-19” OR “coronavirus” AND “care home” OR “nursing home” OR “long term care facility” with no date or language restrictions. We did not identify any publications that focussed on risk of reinfection in seropositive individuals, but subsequent to our search one study has been published using data from two LTCFs in London, UK. This study reported a 96% reduction in the odds of reinfection in individuals who were seropositive compared to those who were seronegative based on 4-month follow-up in 161 participants. We found 10 studies that performed seroprevalence surveys in either staff or staff and residents in LTCFs in 8 cohorts. Five of these were carried out in response to SARS-CoV-2 outbreaks within the care homes, either as part of the subsequent investigation or as post-infection surveillance. The largest of these, which enrolled both staff and residents, was performed in 6 LTCFs and performed longitudinal antibody testing.

*Added value of this study:* We undertook a cohort study in staff and residents from 100 LTCFs in England to investigate whether individuals with evidence of prior SARS-CoV-2 infection could be infected twice. Staff and residents were offered up to three rounds of antibody testing and antibody results were linked to PCR test results which were obtained weekly from staff and monthly from residents through the national SARS-CoV-2 testing programme. This study, which was conducted in >2000 staff and residents, suggests that antibodies provide high levels of protection against reinfection for up to 10 months. Almost all cases of reinfection were symptomatic, but no cases required hospital treatment. Amongst those with detectable baseline antibodies, quantitative antibody titres against spike protein and nucleocapsid were comparable between cases of reinfection and those who did not become reinfected.

*Implications of all available evidence:* Despite high background rates of infection in LTCFs, the overall risk of reinfection was low in this population. This is broadly consistent with findings from large cohort studies of hospital staff, but, importantly, extends the evidence of substantial protection to frail elderly, who are vulnerable to severe outcomes of SARS-CoV-2 due to age-related changes in immunity (immune-senescence) and high levels of comorbidity. The low risk of reinfection in our study suggests identification of immune correlates of protection in this population will require pooling of data across multiple cohorts. As vaccination coverage in residents approaches 100% in England, it will be important to understand whether vaccination and natural infection provide comparable levels of protection against infection. Such insights will inform future policy decisions regarding re-vaccination schedules in LTCF, and the longer-term need for non-pharmaceutical interventions to prevent SARS-CoV-2 transmission, such as asymptomatic testing and visitor restrictions.

## Background

Residents of Long-Term Care Facilities (LTCF) that provide residential and/or nursing care to older people have experienced the highest burden of COVID-19 related mortality of any population group. Older adults may exhibit less robust immune responses to infection due to age-related immune-senescence and underlying co-morbidities, and although emerging data suggest most LTCF residents have a detectable immune response following natural infection with SARS-CoV-2, ^1–4^ the extent to which this protects against a second infection is uncertain. Understanding the degree of protection afforded by prior infection, its duration, and whether primary infection and reinfection differ with regard to disease severity and clinical presentation has major implications for vaccination and for policy decisions regarding the ongoing need or non-pharmaceutical interventions (NPIs) in LTCFs to prevent transmission.

Most individuals who are infected with SARS-CoV-2 develop antibodies against the spike (S) protein and nucleocapsid (N) at 1-2 weeks following symptom onset, ^5^ however data from older age-groups and residents of LTCFs are limited by small sample size. ^3,4^ Neutralising antibodies against the spike protein receptor binding domain (RBD) have been shown to correlate with post-infection immunity, to be dependent on disease severity, ^6^ and to decline over time, ^7^ but understanding of the immune correlates of protection against reinfection remains limited.

Despite the large number of primary infections that have been reported worldwide, there have been relatively few examples of reinfection. ^8–10^ Longitudinal studies in hospital staff suggest reinfections are uncommon, but it is uncertain whether these findings are generalizable to people who live and work in LTCFs due to fundamental differences in underlying health status, age, socio-economic background and levels of exposure to SARS-CoV-2 in each setting.

An estimated 410,000 older people currently live in approximately 11,000 LTCFs in England. ^11^ We undertook a prospective longitudinal cohort study in 100 LTCFs to estimate the incidence and relative hazards of PCR-positive SARS-CoV-2 infection in LTCF staff and residents who were seropositive for SARS-CoV-2 and compared this to staff and residents who were seronegative.

## Methods

The VIVALDI study is a prospective cohort study of staff and residents in LTCFs in England, ^12^ which was established in May 2020 in LTCFs that are owned by the Four Seasons Healthcare Group (FSHCG) and has since expanded to other LTCFs. Participants are being followed up for up to 18 months. This analysis includes data from LTCFs which are owned by the FSHCG. Since June 2020 all staff and residents in LTCFs in England have been offered regular testing for SARS-CoV-2 based on PCR of clinical isolates from nasopharyngeal swabs. ^13^ Residents are tested monthly and staff are tested weekly, although individuals who test positive are then not re-tested for 90 days. ^14^ Local public health teams also investigate outbreaks in LTCFs and usually PCR test all staff and residents at baseline and 7 days later. PCR results (including tests undertaken in hospital) are stored in the COVID-19 Datastore (https://data.england.nhs.uk/covid-19/), which was established as part of the UK’s pandemic response.

Eligible LTCFs were identified by the FSHC and written informed consent to participate was sought from all participants. If residents lacked capacity to consent, a personal or nominated consultee was identified to act on their behalf. Demographic data comprising age, sex, address and whether the individual was a staff member or resident was obtained for all participants. We also retrieved data on symptoms in the 7 days before and after the date of the PCR swab for cases of reinfection, using daily logs of symptoms in staff and residents that were recorded by the FSHCG from March 2020 onwards. Blood sampling was offered to all participants at three time points separated by 6-8 week intervals in June, August and October 2020. Participants could join the study at any blood testing round. Cycle threshold (Ct) values, providing an estimate of viral load, were retrieved for reinfected cases.

### Laboratory methods

Blood samples were tested for IgG to nucleocapsid (N) protein using the Abbott ARCHITECT i system (Abbott, Maidenhead, UK), a semi-quantitative chemiluminescent microparticle immunoassay. Quantitative IgG antibody titres were measured against spike (S) protein and nucleocapsid protein (N) using the MSD V-PLEX COVID-19 Respiratory Panel 2 (96-well, 10 Spot Plate was coated with three SARS CoV-2 antigens (S, S-RBD S-NTD and N)) (Cat # K15372U) from Meso Scale Diagnostics, Rockville, MD USA (Appendix). PCR samples were tested in a network of laboratories using a range of assays which targeted different genes (appendix pp 2).

We used an index value cut-off of 0·8 to classify samples as antibody positive (≥ 0·8) or antibody negative (<0·8) to maximise the sensitivity of the Abbott assay whilst maintaining high specificity. ^15,16^ Quantitative IgG antibody titres were obtained for cases of suspected reinfection and compared to control samples from participants who met the following criteria: no record of PCR positive test during the study; antibody positive at baseline; at least one PCR negative test during follow-up.

### Data Linkage

Approximately 60% of PCR results from the national testing programme, and almost all tests undertaken in hospital, can be linked to staff and residents using a pseudo-identifier which is based on the individuals’ unique National Health Service (NHS) number. PCR results from the national testing programme are also linked to specific care homes using the Care Quality Commission’s unique location ID (CQC-ID). The Care Quality Commission regulates all providers of health and social care in England.

Antibody test results were submitted to NHS England and matched to NHS number using an algorithm based on participant forename and surname, date of birth, sex and postcode. This made it possible to generate a common pseudo-identifier to link antibody and PCR test results in the COVID-19 Datastore. Dates of first vaccination against SARS-CoV-2 were retrieved for all participants through linkage to the National Immunisation Database, based on the same pseudo-identifier. Analysis was undertaken in the UCL Data Safe Haven.

### Inclusion criteria, exposures and start of time at risk

Staff and residents were eligible for inclusion in the analysis of risk of re-infection if they had a valid pseudo-identifier (enabling linkage of antibody test results to PCR tests); they lived or worked in a LTCF that was owned by FSHCG; they had at least one PCR test result during the analysis period; and they had at least one antibody test during the study period. We excluded staff with a recorded age > 65 years and residents aged < 65 years.

All participants were classified into two cohorts (positive and negative) according to their first (baseline) antibody test. Exposure status was based on IgG antibodies to nucleocapsid (Abbott) because this test was available for all participants. Subsequent seroconversion was not considered in our primary analysis due to small numbers of participants in which this occurred (see Results section).

The time-at-risk ‘entry time’ for participants was 1^st^ October 2020 or 28 days after their first available antibody test, whichever was later. 91% joined the study on October 1st. These restrictions reduced the risk of misclassifying prolonged PCR positivity as reinfection, particularly between July and September 2020 when the incidence of SARS-CoV-2 was comparatively low in England. ^17,18^ For example, prior to October we identified 13 and 7 infections among antibody-positive and antibody-negative participants respectively, suggesting a high risk of misclassification in this period.

### Primary outcome and end of time at risk

All positive PCR tests after entry time were considered to indicate infection or reinfection. Participants were followed up from entry time until the earliest of the following dates: first positive PCR test (main ‘failure’ outcome); last PCR test (removes individuals who have left the LTCF as most staff and residents undergo regular PCR testing); 12 days following the first vaccination of any resident in the home (residents); 12 days following the first vaccination of any staff member in the home (staff). The 12-day window was chosen based on evidence that the protective effect of vaccination begins after 12 days. ^19^ The date of first vaccination in the care home was preferred over the date each individual was vaccinated because vaccination records may be incomplete, and most Facilities achieved high vaccine coverage.

Quantitative antibody titres to spike and nucleocapsid were retrieved for all cases of suspected reinfection. For comparison, control samples were retrieved for 23 residents and 19 staff from five randomly selected care homes who met the following criteria: antibodies to nucleocapsid (Abbott) detected at the first blood testing round; three antibody tests; no record of PCR positive test; at least one PCR test result during the analysis period.

### Statistical Analysis methods

Kaplan Meier curves are presented separately for staff and residents, illustrating the cumulative probability of testing PCR positive over time by baseline antibody status. These curves exclude late entrants to the analysis (n=165) to allow presentation on a calendar time scale beginning on 1st October 2020.

Cox regression was used to estimate hazard ratios (HRs) for baseline antibody positivity. The baseline hazard was defined over calendar time, with participants entering the ‘risk set’ on their entry date (in most cases 1^st^ October 2020). Our primary analysis is ‘within LTCF’ to remove potential confounding from unmeasured LTCF factors, different trends in background incidence, or differences in the proportion of individuals in each LTCF who had experienced prior infection. This was based on Cox regression stratified by LTCF to allow a different baseline hazard for each LTCF, restricted to LTCFs that had participants with and without baseline antibodies and also some positive PCR tests after study entry. An alternative stratification by regions of England (i.e., those in Table 1) was also conducted, which whilst more susceptible to bias leads to greater precision. In this model, confidence intervals for the HRs are presented based on robust standard errors to acknowledge the clustering of participants by LTCF. For both models, adjusted hazard ratios were estimated, adjusting for gender and for the non-linear effects of age using cubic splines with 5 knots at default positions.

**Table 1:** Baseline characteristics of participants by baseline antibody status and subject type (n=2111)

|  | All residents | Residents negative | Residents positive | All staff | Staff negative | Staff positive |
| --- | --- | --- | --- | --- | --- | --- |
| **TOTAL** |  |  |  |  |  |  |
| n | 682 | 456 | 226 | 1429 | 1021 | 408 |
| **Age** |  |  |  |  |  |  |
| Median [IQR] | 86 [79-91] | 86 [80-92] | 86 [79-91] | 47 [34-56] | 46 [33-56] | 48 [36-57] |
| Range | 65-103 | 65-102 | 65-103 | 18-65 | 18-65 | 20-65 |
| **Gender - n (%)** |  |  |  |  |  |  |
| Male | 208 (30.5) | 137 (30.0) | 71 (31.4) | 174 (12.2) | 122 (12.0) | 52 (12.8) |
| Female | 474 (69.5) | 319 (70.0) | 155 (68.6) | 1,255 (87.8) | 899 (88.1) | 356 (87.3) |
| **Region - n (%)** |  |  |  |  |  |  |
| East Midlands | 66 (9.7) | 58 (12.7) | 8 (3.5) | 136 (9.5) | 117 (11.5) | 19 (4.7) |
| Eastern | 78 (11.4) | 62 (13.6) | 16 (7.1) | 77 (5.4) | 70 (6.9) | 7 (1.7) |
| London | 96 (14.1) | 53 (11.6) | 43 (19.0) | 120 (8.4) | 71 (7.0) | 49 (12.0) |
| North East | 128 (18.8) | 60 (13.2) | 68 (30.1) | 355 (24.8) | 217 (21.3) | 138 (33.8) |
| North West | 64 (9.4) | 34 (7.5) | 30 (13.3) | 265 (18.5) | 173 (16.9) | 92 (22.6) |
| South East | 78 (11.4) | 44 (9.7) | 34 (15.0) | 129 (9.0) | 83 (8.1) | 46 (11.3) |
| South West | 34 (5.0) | 30 (6.6) | 4 (1.8) | 55 (3.9) | 49 (4.8) | 6 (1.5) |
| West Midlands | 61 (8.9) | 51 (11.2) | 10 (4.4) | 108 (7.6) | 86 (8.4) | 22 (5.4) |
| Yorkshire & Humber | 77 (11.3) | 64 (14.0) | 13 (5.8) | 184 (12.9) | 155 (15.2) | 29 (7.1) |
| **Total number of PCR tests (June 2020 - February 2021)** |  |  |  |  |  |  |
| Median [IQR] | 7 [5-9] | 7 [5-9] | 7 [5-10] | 17 [9-26] | 16 [9-24] | 20 [11-29] |
| Range | 1-18 | 1-17 | 1-18 | 1-55 | 1-55 | 1-52 |
| **Number of PCR tests during analysis time at risk** |  |  |  |  |  |  |
| Median [IQR] | 3 [2-4] | 3 [2-5] | 3 [2-4] | 7 [4-11] | 7 [4-11] | 8 [5-11] |
| Range | 1-11 | 1-11 | 1-10 | 1-22 | 1-21 | 1-22 |
| **Total available antibody test results - n (%)** |  |  |  |  |  |  |
| 1 | 117 (17.2) | 86 (18.9) | 31 (13.7) | 510 (35.7) | 385 (37.7) | 125 (30.6) |
| 2 | 173 (25.4) | 121 (26.5) | 52 (23.0) | 457 (32.0) | 337 (33.0) | 120 (29.4) |
| 3 | 392 (57.5) | 249 (54.6) | 143 (63.3) | 462 (32.3) | 299 (29.3) | 163 (40.0) |

Quantitative antibody titres were compared between reinfection cases and controls (Appendix pp 2). Sensitivity analysis was undertaken to investigate the impact of using the manufacturer’s index value threshold (1.4) for the Abbott assay (Appendix Sensitivity Analysis 1), and the impact of including analysis time prior to 1st October 2020 (Appendix Sensitivity Analysis 2).

Sample size for the VIVALDI study was based on the precision of estimates for antibody prevalence. ^12^

All analysis was conducted in STATA v16.0.

### Ethics

Ethical approval for this study was obtained from the South Central - Hampshire B Research Ethics Committee, REC Ref: 20/SC/0238.

## Results

A total of 2111 participants, comprising 682 residents and 1429 staff, met the study inclusion criteria (Table 1, Appendix Figure S1). The mean number of residents and staff in each LTCF was 6·8 (SD 6·9) and 14·3 (SD 10·0) respectively. The cohort was predominantly female, both among residents (n=474; 70%) and staff (n=1255; 88%). Baseline antibodies to nucleocapsid were detected in 226 residents (33%) and 408 staff (29%). The median age of residents was 86 years (Interquartile range, IQR: 79-91) and 47 years in staff (IQR: 34-56). Participants from all regions of England were represented in the sample.

392 of 682 residents (58%) and 462 of 1429 (32%) of staff participated in all three rounds of blood testing. Around 2% of both residents (10/456) and staff (19/1021) who tested negative for antibodies against nucleocapsid in their first testing round subsequently had a positive antibody test. Conversely, around 17% of residents (39/226) and 25% of staff (102/408) who had antibodies against nucleocapsid at baseline tested antibody-negative in a later round. Residents and staff had a median of 3 (IQR: 2-4) and 7 (IQR: 4-11) PCR tests during the analysis-time respectively.

Staff and residents contributed 3749 and 1809 months of follow-up time respectively, Table 2. A total of 93 antibody negative residents had a PCR positive test (0·054 per month at risk) compared to 4 antibody positive residents (0·007 per month at risk). There were 111 PCR positive tests in antibody negative staff (0·042 per month at risk) compared to 10 in antibody positive staff (0·009 per month at risk)

**Table 2:** Total number of participants, infections and time at risk by baseline antibody status and participant type.

|  | Number of participants | Person-time at risk (months) | Number of PCR positive infections (%) | Rate of PCR positive infection per month at risk |
| --- | --- | --- | --- | --- |
| **Residents** | 682 | 1,809 | 97 (14.2) | 0.054 |
| First antibody -ve | 456 | 1203 | 93 (20.4) | 0.077 |
| First antibody +ve | 226 | 606 | 4 (1.8) | 0.007 |
| **Staff** | 1,429 | 3,749 | 121 (8.5) | 0.032 |
| First antibody -ve | 1,021 | 2,663 | 111 (10.9) | 0.042 |
| First antibody +ve | 408 | 1,086 | 10 (2.5) | 0.009 |

Figure 1 shows the proportion of residents and staff who remain PCR test negative during the analysis time by antibody status at baseline. The incidence of infection gradually increases over time in staff and residents, and the curves indicate low numbers of infection in individuals who are antibody positive. The difference between antibody positive and negative curves is greater for residents than for staff. By the end of January 2021, very few participants remain at-risk in the analysis, due to the rapid roll-out of vaccination in LTCFs in England from December 2020.

**Figure 1.**
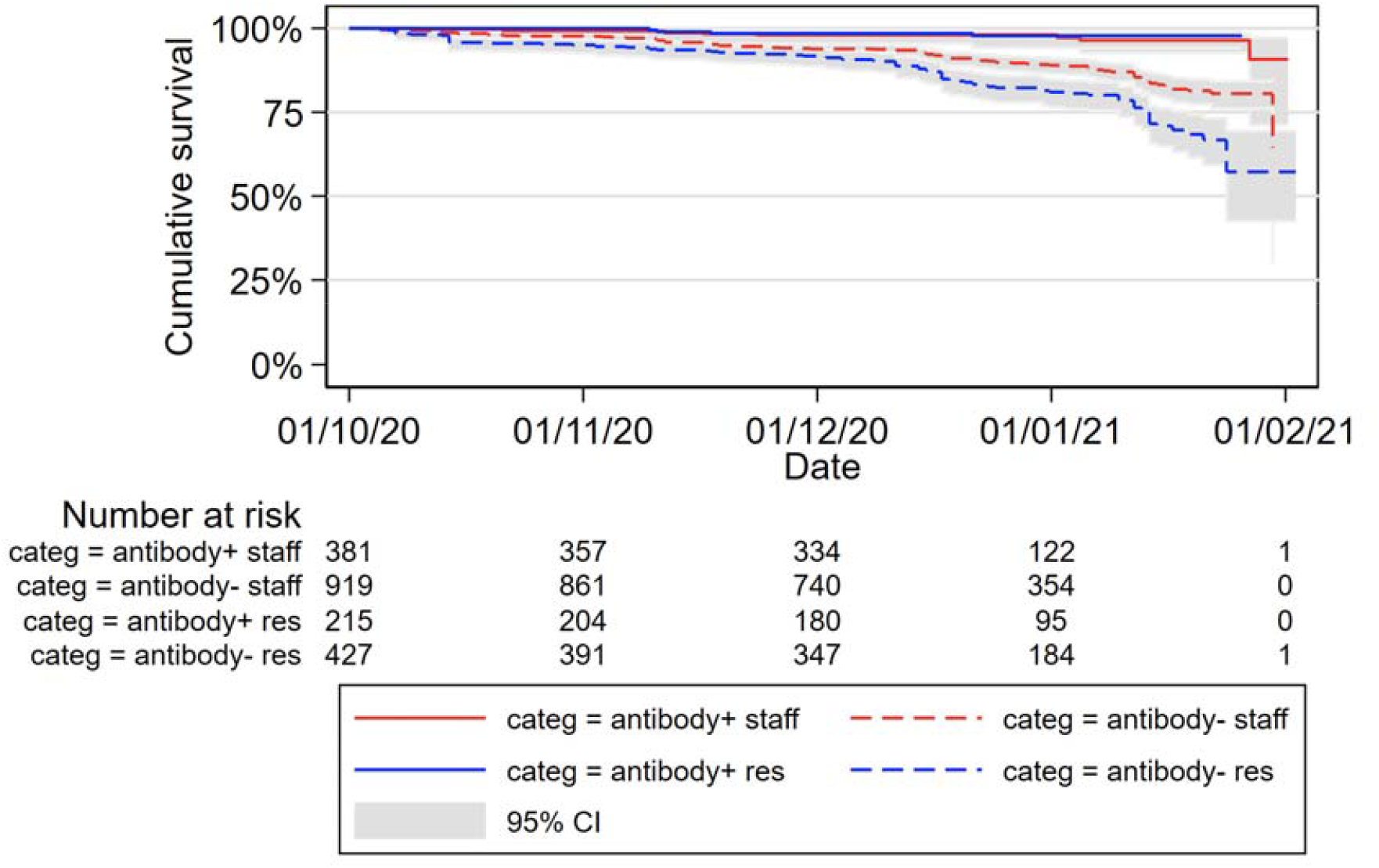
Kaplan-Meier estimates of PCR positive infections by baseline antibody status and participant type. +/-refer to outcome of baseline antibody test for each participant. Kaplan-Meier estimates exclude late entrants to the analysis (n=165) to allow presentation on a calendar time scale beginning on 1st October 2020.

In the Cox regression stratified by LTCF, the relative adjusted hazard ratios for PCR positive infection were 0·15 (0·05-0·44) and 0·39 (0·19-0·82) comparing seropositive versus seronegative residents and staff respectively, Table 3. The estimated protective effect is slightly stronger when stratifying by region, but this analysis may be subject to some confounding.

**Table 3:** Multivariate analysis of risk of PCR positive infection by baseline antibody status, stratified by LTCF or by region.

|  | Stratified by LTCF |  | Stratified by region |  |
| --- | --- | --- | --- | --- |
|  | aHR [95% CI] | p-value | aHR [95% CI] | p-value |
| **Residents** | 0.15 [0.05-0.44] | p=0.0006 | 0.08 [0.03-0.23] | p<0.0001 |
| **Staff** | 0.39 [0.19-0.82] | p=0.0123 | 0.26 [0.12-0.54] | p=0.0003 |

*aHR: hazard ratio antibody positive vs. negative, adjusted for age and gender*

Information on whether the resident or staff member had symptoms in the 7 days before or after PCR testing was available for 12 of 14 cases of reinfection, Appendix Table S1. All residents (4/4) with reinfection were febrile at or around the time of their PCR test and of 8 staff members, 6 reported cough, 1 reported fever and 1 was asymptomatic. By comparison, 14/42 controls had symptoms in the 7 days before or after their PCR test. None of the reinfection cases were admitted to hospital or died as a result of their infection and the median duration of symptoms was 7 (5-13) days.

Ct values were retrieved for 13/14 reinfection samples. The median Ct value for reinfection cases was 36 (30·1-37·0). 6/7 samples that were analysed using the same PCR assay, and 9/14 samples that were tested using assays that targeted the ORF1ab had Ct values >30 (Appendix Table S1).

Quantitative antibody data were available for 11 of the 14 reinfection cases, and 42 control participants who were antibody positive at baseline and remained PCR negative throughout follow-up. There was no statistically significant difference in antibody titres to spike and nucleocapsid in individuals who were re-infected and those who remained PCR-negative during follow-up, when considering antibodies at the first testing round (baseline), and at the last antibody testing round stratified by the time gap between the antibody test and the PCR test, Figures 2 & 3 and Appendix Figure S2.

**Figure 2.**
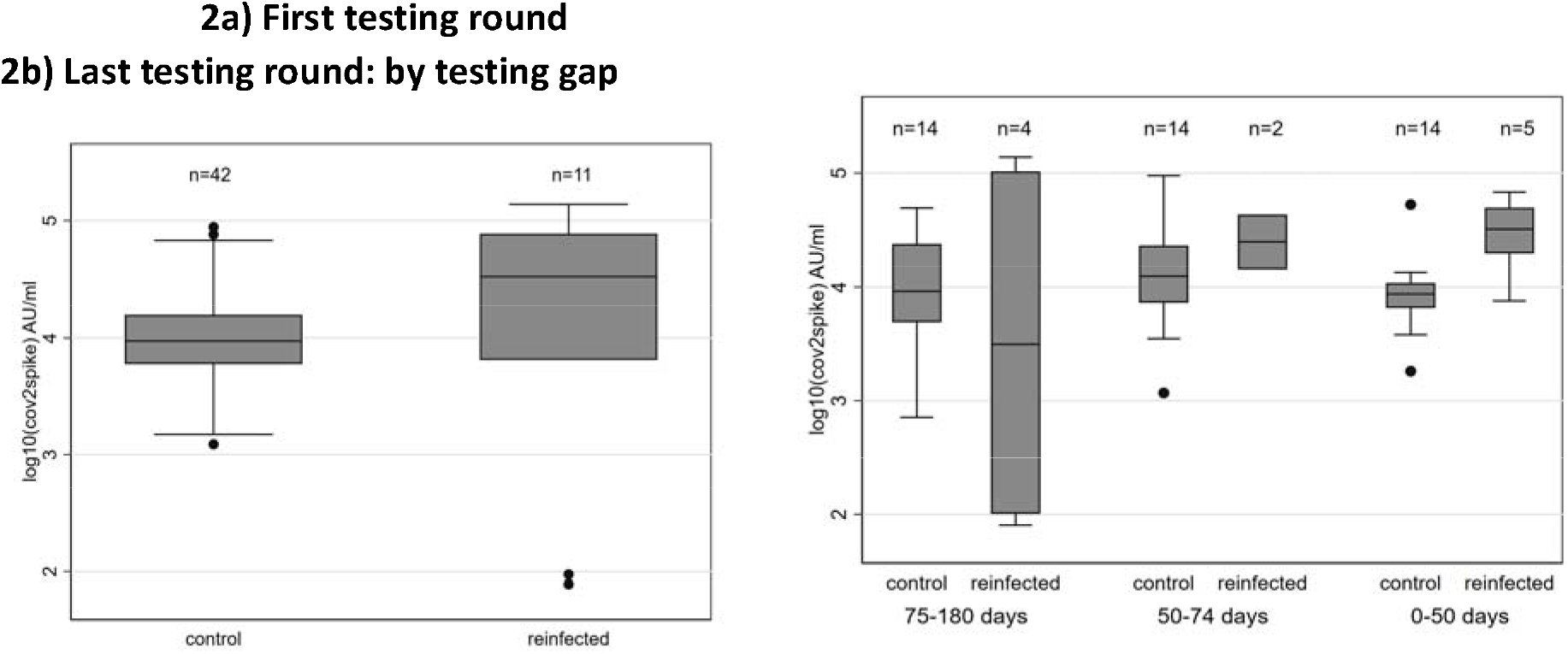
Quantitative SARS-CoV-2-spike IgG titre values by reinfection status by a) first testing round and b) last testing round by testing gap. Figure 2 compares quantitative spike values on a logarithmic scale by reinfection status. Median baseline spike levels are 33,057 AU/ml for reinfection cases [IQR 6468-77315] compared to 9,457 AU/ml in control cases [IQR 5978-15820]. The median gap between the last antibody test and last relevant PCR result (first positive for cases, last negative for controls) was 62 days for reinfection cases [IQR 28-88] and 68 days for controls [IQR 48-75]. Based on this testing gap, participants were categorised into three categories: 0-50 days between tests, 50-75 days, and 75-180 days (see Figure 2b). Differences in levels of antibodies to spike between reinfection cases and controls are not statistically significant for either the first testing round (p=0·751) or for the last available antibody test when controlling for testing gap (p=0·762).

**Figure 3.**
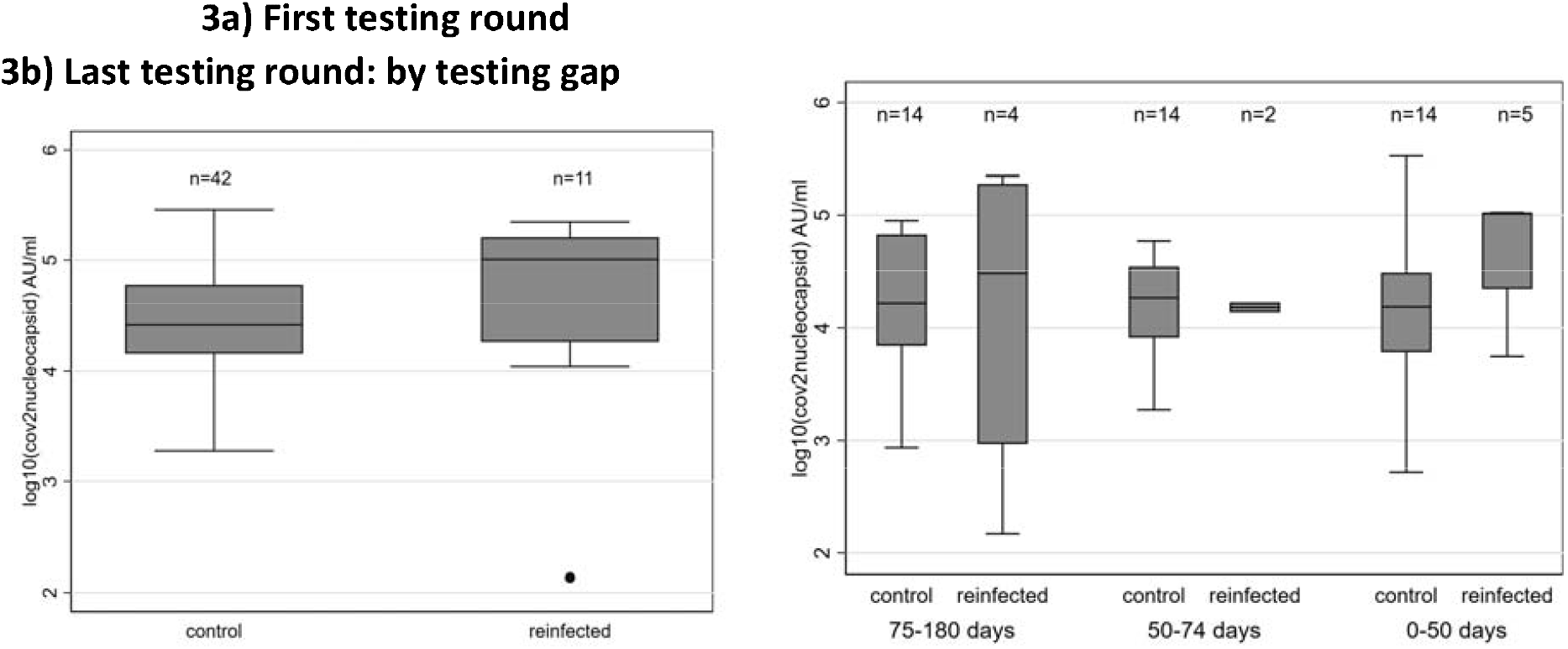
Quantitative SARS-CoV-2-nucleocapsid IgG titre values by reinfection status by a) first round of testing and b) last round of testing. Figure 3 compares quantitative nucleocapsid values on a logarithmic scale by reinfection status. Median baseline levels of antibodies to nucleocapsid are 101,527 AU/ml for reinfection cases [IQR 18393-161580] compared to 26,326 AU/ml in control cases [IQR 14378-59633]. Again, differences in log10 levels of antibodies to nucleocapsid are not statistically significant for either the first testing round (p=0·544) or for the last available antibody test when controlling for testing gap (p=0·426).

Sensitivity analyses using the manufacturer’s recommended index cut off value (1.4) for the Abbott test and assuming an entry date of 28 days following the first antibody test for all participants did not substantially alter our findings (Appendix Table S2).

## Discussion

In this cohort study in 100 LTCFs, the risk of PCR positive SARS-CoV-2 infection was substantially lower in residents and staff with baseline SARS-CoV-2-specific antibodies. We detected only 14 cases of possible reinfection, mainly affecting staff, and although almost all these cases reported symptoms, none required hospital treatment for a reinfection. This suggests that prior infection provides a high degree of protection against a second infection and is broadly consistent with findings from longitudinal studies in hospital staff. Whilst staff and residents with baseline antibodies to nucleocapsid remain vulnerable to symptomatic infection, our findings based on up to 10 months follow-up from primary infection (assuming earliest infections occurred in March 2020) suggest that their risk of reinfection is low (< 1% per month). Similar findings were obtained in sensitivity analyses which varied the threshold for detection of IgG to nucleocapsid, and the person-time at risk.

The low number of reinfections limited statistical power to draw strong conclusions about symptom profiles, but it was notable that the majority of reinfections in both staff and residents were symptomatic. This contrasts with studies of hospital staff, in which one-third of reinfected cases reported symptoms. ^17^ Symptoms tended to be milder in staff (cough) compared to residents (fever), but we cannot rule out differential ascertainment of symptoms between staff and residents. The risk of recall bias is minimised by the fact that LTCFs recorded symptoms in staff and residents prospectively.

We retrieved Ct values for reinfection samples but were unable to compare viral load between individual’s reinfections and primary infections due to lack of testing at the start of the pandemic. In the subset of reinfection samples that were tested using comparable PCR assays, the majority (64-86%) of samples had Ct values >30 - with Ct >30 being a widely used threshold to denote lower viral load. ^20,21^ By comparison, >80% of samples obtained from PCR testing in LTCFs over the same time period, which will mainly represent primary infections given the rarity of reinfection, had Ct values <30, based on a comparable assay. ^22^ Although it is difficult to compare tests conducted in different laboratories, this provides tentative evidence that reinfections may be associated with lower viral load and reduced likelihood of transmission compared to primary infections. Ideally, the presence of reinfection would have been confirmed by viral sequencing. Nevertheless, it is unlikely that we misclassified primary infections that remained PCR-positive as reinfections, because most participants had at least 90 days and all had 2 or more negative PCR tests between their baseline antibody test and PCR positive test.

We found no difference in quantitative antibody titres against spike protein or nucleocapsid in reinfected cases compared to uninfected cases with baseline antibodies. The rarity of reinfection makes studies of correlates of protection challenging and highlights the need to standardise the assays that are used to evaluate humoral and cellular immunity so sample collections and results can be pooled across cohorts. ^15^ As we did not know the date of primary infection for most reinfected cases in our study (due to low levels of PCR testing in LTCFs at the start of the pandemic), it is possible that these cases followed primary infections that occurred at the start of the pandemic and/or were asymptomatic associated with low viral load. The reasons why the magnitude of protection against reinfection afforded by baseline antibodies was greater in staff than residents are unclear, but a possible explanation is that symptomatic staff accessed PCR testing outside of the LTCF.

A strength of our study is that we estimated the incidence of infection during a period of high community prevalence of SARS-CoV-2 in the UK, associated with the rapid emergence of the B.1.1.7 variant. ^21^ The local prevalence of SARS-CoV-2 is likely to be a key driver of transmission in LTCFs because staff can import infection from the community. ^23,24^ Restricting our analysis to October 1st onwards reduced the amount of ‘person-time at risk’ to 4 months, though incidence was very low in the preceding months. As most participants already had baseline antibodies on 1st October it is likely that they were mainly infected during the first wave of the pandemic, up to 6 months earlier.

Our study is limited by sample size and the quality of surveillance data. ^25,26^ We recruited a median of 6·8 residents and 14·3 staff per LTCF, reflecting the challenges of recruiting frail residents who may lack capacity to consent and/or be receiving end of life care. During the period of analysis, residents (who are tested monthly) and staff (who are tested weekly) had an average of 1·6 and 3·1 PCR tests per month respectively, suggesting high levels of participation in the voluntary testing programme. Less frequent PCR testing in residents and missing test results for staff will tend to underestimate rates of asymptomatic infection in both groups. However, it is likely that the majority of infections from June 2020 onwards were detected because most participants were tested at least every 4 weeks, and the median duration of PCR positivity is 12-18 days. ^27,28^ We classified baseline antibody status against nucleocapsid because data on antibodies to spike were only available for a subset of participants, and this may have underestimated the proportion of individuals with previous infection due to antibody waning. To overcome this limitation, we reduced the test index threshold for the Abbott assay to 0.8, which has been shown to increase sensitivity with minimal effect on test specificity.

As vaccination coverage in residents approaches 100%, ^29^ it will be important to understand whether vaccination and natural infection provide comparable levels of protection against new infection. New infections were still evident in our dataset in February 2021, and work is ongoing to investigate the effectiveness of different vaccine types and dosing schedules in this population. However, the high degree of vaccine coverage in residents will make it challenging to investigate protective effectiveness in this group.

In summary, the risk of a second SARS-CoV-2 infection is substantially reduced in staff and residents of LTCFs who are SARS-CoV-2 seropositive and the observed reinfections were not clinically severe. Understanding the correlates of immunity that protect against future infection will be fundamental to policy decisions regarding LTCFs, including re-vaccination schedules and the ongoing need for NPIs to prevent SARS-CoV-2 transmission.

## Data Availability

De-identified test results and limited meta-data will be made available for use by researchers in future studies, subject to appropriate research ethical approvals, once the VIVALDI study cohort has been finalised. These datasets will be accessible via the Health Data Research UK Gateway https://www.healthdatagateway.org

## Contributors

Study conceptualisation: LS, AH, AC, TP and MK; Statistical methodology: AC, TP, LS, AH, MK; Formal analysis: AC, TP; Project administration: MK, CF, JR, MS and AIS; Data curation and validation: HW, DD; Literature review: MK; Funding: LS, AH, AC; Laboratory investigations: GT, PM; Writing (original draft): LS, TP, MK; Writing (review and editing): MK, TP, GT, CF, HW, AIS, MS, DD, JR, AH, PM, AC, LS. TP, AC, LS and MK had full access to the data in the study. LS and AC have shared responsibility for the decision to submit for publication.

## Data Sharing

De-identified test results and limited meta-data will be made available for use by researchers in future studies, subject to appropriate research ethical approvals, once the VIVALDI study cohort has been finalised. These datasets will be accessible via the Health Data Research UK Gateway https://www.healthdatagateway.org/.

## Acknowledgements

The authors would like to thank the staff and residents in the LTCFs that participated in this study, and Mark Marshall at NHS England who pseudonymised the electronic health records. This report is independent research funded by the Department of Health and Social Care (COVID-19 surveillance studies). AH is supported by Health Data Research UK (HDR-UK; grant no LOND1), which is funded by the UK Medical Research Council, Engineering and Physical Sciences Research Council, Economic and Social Research Council, Department of Health and Social Care (England), Chief Scientist Office of the Scottish Government Health and Social Care Directorates, Health and Social Care Research and Development Division (Welsh Government), Public Health Agency (Northern Ireland), British Heart Foundation, and Wellcome Trust. MK is funded by a Wellcome Trust Clinical PhD Fellowship. LS is funded by a National Institute for Health Research (NIHR) Clinician Scientist Award (CS-2016-007). The views expressed in this publication are those of the authors and not necessarily those of the NHS, Public Health England, or the Department of Health and Social Care.

