## Supplementary Tables and Methods for "Incidence of SARS-CoV-2 infection according to baseline antibody status in staff and residents of 100 Long Term Care Facilities (VIVALDI study)"

### **Description of the Cohort**

**Figure S1. Study flow diagram**

4470 participants undergo ≥1 round of antibody testing in 100 LTCFs

1081 residents; 3389 staff

1575 participants (181 residents, 1394 staff) excluded that could not be linked to a NHS identifier.

2895 results could be linked to a NHS identifier

900 residents

1995 staff

114 participants (42 residents, 72 staff) excluded based on age cut-offs (> 65 years for staff, < 65 years for residents).

2781 have appropriate age for subject type

858 residents

1923 staff

501 participants excluded (154 residents, 347 staff) with zero PCR tests after 1^st^ October 2020.

2280 ≥1 PCR test result after entry date (01/10/20)

704 residents

1576 staff

169 participants (22 residents, 147 staff) excluded as no PCR test before vaccination censor date.

2111 participants with ≥1 PCR test result before vaccination censor date

682 residents from 86 LTCFs

1429 staff from 97 LTCFs

### **MSD Assay detailed protocol**

MSD V-PLEX COVID-19 Respiratory Panel 2 (Cat # K15372U) from Meso Scale Diagnostics, Rockville, MD USA Antigens were spotted at 200−400 μg/mL. Multiplex MSD Assays were performed as per the instructions of the manufacturer. To measure IgG antibodies, 96-well plates were blocked with MSD Blocker A for 30 minutes. Following washing with washing buffer, samples diluted 1:500 in diluent buffer, as well as the reference standard and internal controls were added to the wells. After 2-hour incubation and a washing step, detection antibody (MSD SULFO-TAG™ Anti-Human IgG Antibody, 1/200) was added. Following washing, MSD GOLD™ Read Buffer B was added and plates were read using a MESO ® SECTOR S 600 Reader. Text files from the machine were then read on MSD Discovery Workbench and data exported as .csv files. The values from exported data were then adjusted for any sample dilution. Assay Cut-offs were determined by running pre-pandemic plasma samples from healthy donors on the same platform. Cut-offs used are Spike 350 AU/ml and Nucleocapsid 1200 AU/ml.

### **Polymerase Chain Reaction (PCR) Testing**

Pillar 1, which comprises samples from public health led outbreak investigations and all tests undertaken in hospital; and Pillar 2, which processes samples from community testing programmes in settings such as LTCFs, educational settings and mobile testing centres. Pillar 2 community testing was not widely available in LTCFs until June 2020. Pillar 1 samples from this study were processed by each local NHS diagnostic laboratory (66 laboratories). Pillar 2 samples were processed by a national network of 59 accredited laboratories which was established during the pandemic to provide rapid PCR testing at scale. A range of PCR assays targeting different genes were used across these laboratories. Details of the assays used to analyse samples from reinfections are included in Supplementary Table S1.

### **Statistical analysis of quantitative antibody titres**

Quantitative antibody titres at the first (baseline) and last testing rounds were summarised (Figures 2 and 3 in the main manuscript), the latter stratified by the testing gap between the antibody test and the ‘relevant’ PCR test (first positive for reinfection cases and last negative for controls). Note all reinfections occurred after the last antibody test. Differences in baseline and last antibody test results between reinfections and controls were tested using separate linear regressions of log10 antibody titre. Both models accounted for clustering within LTCFs by using robust standard errors. In the model for last antibody test, the potentially non-linear impact of the testing gap was represented using cubic splines with 5 knots at default positions.

### **Quantitative antibody titres against nucleocapsid and spike protein**

Figure S2 shows quantitative values for antibodies to both nucleocapsid and spike on a logarithmic scale which were measured in up to 3 blood testing rounds undertaken in June/July, August/September and October/November 2020. Of 11 reinfection cases, 8 had > 1 antibody test during follow-up.

**Figure S2. Longitudinal quantitative IgG titres against a) nucleocapsid and b) spike in 11 cases of reinfection by testing round.**

**a) SARS-CoV-2-nucleocapsid                                                 b) SARS-CoV-2-spike**

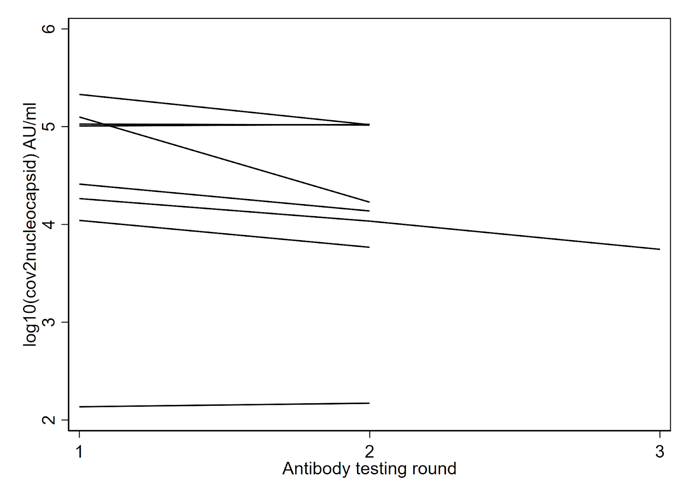

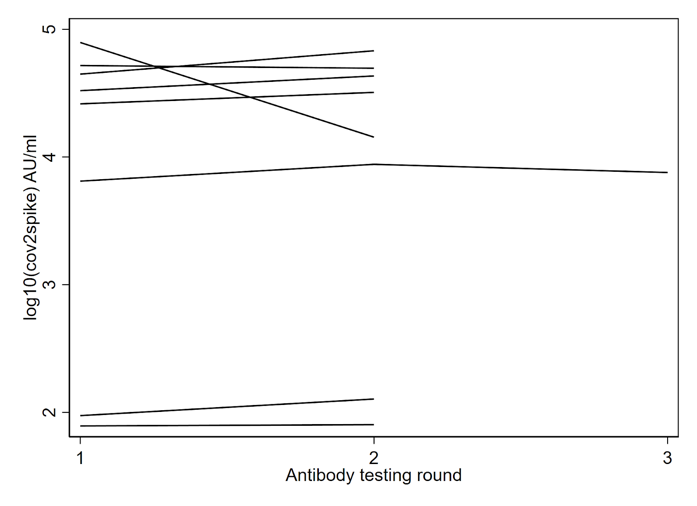

### **Description of reinfection cases**

Results of PCR and antibody testing were obtained where available for 14 reinfection cases along with detailed data on symptoms. PCR Cycle threshold (Ct) values were retrieved for positive samples taken at the time of reinfection. Samples were processed in four Pillar 2 laboratories using different assays however all targeted at least the ORF1ab gene. Antibody testing was performed at one to three time points for all samples using the Abbott assay on all samples and the MSD assay where samples were available. Results and details of PCR assays performed are presented in Table S1.

**Table S1. Characteristics of staff and residents with suspected reinfection**

| **Age band / sex, care home role** | **Baseline serology to reinfection (days)** | **Symptoms at reinfection** | **Symptom duration (days)** | **Ct value of reinfection PCR ^~^** | **Negative NP PCR samples between baseline serology and reinfection** | **Antibody results** | | | |
| --- | --- | --- | --- | --- | --- | --- | --- | --- | --- |
|  |  |  |  |  |  | **Month / year of sample (days before reinfection)** | **SARS-CoV-2 spike IgG AU/ml (MSD)** | **SARS-CoV-2 nucleocapsid IgG AU/ml (MSD)** | **SARS-CoV-2 nucleocapsid IgG Index S/C (Abbott)** |
| 70-80 F, R | 116 | Fever | 5 | 30·1 * | 2 | 07/20 (116) | 137840 | 222308 | 6·27 |
|  |  |  |  |  |  | 11/20 (4) | NA | NA | 6·35 |
| 90-100 F, R | 140 | Fever | 7 | 41·4 ^±^ | 4 | 06/20 (140) | NA | NA | 8·69 |
|  |  |  |  |  |  | 08/20 (84) | 26055 | 106219 | 6·77 |
|  |  |  |  |  |  | 10/20 (28) | 32051 | 103738 | 8·48 |
| 60-70 F, R | 188 | Fever | 3 | 28·9 ^±^ | 6 | 06/20 (188) | NA | NA | 1·06 |
|  |  |  |  |  |  | 08/20 (132) | 94 | 10997 | 0·35 |
|  |  |  |  |  |  | 10/20 (76) | 127 | 5839 | 0·18 |
| 70-80M, R | 148 | Fever | 5 | NA | 4 | 06/20 (148) | NA | NA | 1·1 |
|  |  |  |  |  |  | 08/20 (92) | NA | NA | 1·24 |
| 50-60 F, S | 128 | Cough | 7 | 37·7 ^±^ | 9 | 07/20 (128) | 33057 | 25849 | 2·82 |
|  |  |  |  |  |  | 09/20 (71) | 43078 | 13727 | 1·42 |
| 40-50 M, S | 132 | Cough | 10 | 37·6 ^±^ | 13 | 06/20 (132) | NA | NA | 7·63 |
|  |  |  |  |  |  | 08/20 (76) | 51997 | 101527 | 7·77 |
|  |  |  |  |  |  | 10/20 (20) | 49532 | 105247 | 9·07 |
| 50-60 F, S | 103 | Cough | 11 | 35·9 ^#^ | 6 | 06/20 (103) | 77315 | 161580 | 5·5 |
| 50-60 F, S | 200 | Fever | 14 | 25·2 ^#^ | 12 | 06/20 (200) | NA | NA | 2·1 |
|  |  |  |  |  |  | 08/20 (144) | 78 | 137 | 1·63 |
|  |  |  |  |  |  | 10/20 (88) | 80 | 149 | 1·87 |
| 30-40 F, S | 124 | Cough | 14 | 36·3 ^±^ | 9 | 07/20 (124) | 6468 | 18393 | 2·56 |
|  |  |  |  |  |  | 09/20 (68) | 8759 | 10810 | 1·39 |
|  |  |  |  |  |  | 11/20 (12) | 7557 | 5565 | 0·76 |
| 60-70 F, S | 104 | Cough | 14 | 36·9  ^#^ | 9 | 07/20 (104) | 44544 | 213650 | 5·58 |
|  |  |  |  |  |  | 09/20 (48) | 67856 | 104658 | 3·51 |
| 50-60 F, S | 141 | Unknown | - | 35·2  ^#^ | 2 | 08/20 (141) | NA | NA | 7·39 |
| 40-50 F, S | 118 | Cough | 5 | 36·2 ^±^ | 12 | 07/20 (118) | 78784 | 125512 | 1·44 |
|  |  |  |  |  |  | 09/20 (62) | 14292 | 16888 | 0·71 |
|  |  |  |  |  |  | 11/20 (6) | NA | NA | 0·4 |
| 60-70  M, S | 153 | No | n/a | 28·9 * | 13 | 06/20 (153) | NA | NA | 3·72 |
|  |  |  |  |  |  | 08/20 (97) | NA | NA | 2·41 |
|  |  |  |  |  |  | 10/20 (41) | 19580 | 22254 | 1·35 |
| 30-40 F, S | 147 | Unknown | - | 39·6  ^±^ | 23 | 09/20 (23) | NA | NA | 1·37 |

R  Resident         	             	S  Staff	NA Data unavailable

~ If > 1 gene target, mean Ct value presented

*Primer Design PCR assay with ORF1ab gene target, Ct threshold for positivity = 40.

# Randox PCR assay that targets ORF1ab and E genes, Ct threshold for positivity = 37.

± Perkin Elmer SARS-CoV-2 Real-time RT-PCR Assay CE-IVD with two target genes (N and Orf1ab) and one human IC gene (RPP30), Ct threshold for positivity = 4

### **Sensitivity analysis 1**

Table S2 shows the results of repeating the multivariable analysis using the manufacturer’s recommended cut off value of >1.4 to classify samples as positive or negative for SARS-CoV-2 (based on detection of antibodies to nucleocapsid).

**Table S2: Multivariate analysis of risk of infection by antibody status (1.4 Abbott threshold)**

|  | **Stratified by LTCF** | | **Stratified by region** | |
| --- | --- | --- | --- | --- |
|  | aHR [95% CI] | p-value | aHR [95% CI] | p-value |
| **Residents** | 0·08 [0·02-0·35] | p=0·001 | 0·05 [0·01-0·21] | p<0·001 |
| **Staff** | 0·43 [0·20-0·92] | p=0·029 | 0·28 [0·13-0·60] | p=0·001 |

aHR adjusted for age and gender

### **Sensitivity analysis 2**

Table S3-S4 assumes an entry date of 28 days following the first antibody test for all participants, removing the restriction using October 1st as a minimum entry date.

This alternative approach results in a larger sample than in our main analysis (n=2,220). Table SA2.1 presents analysis for our main sample, while Table SA2.2 presents analysis including these additional participants.

**Table S3**: **Multivariate analysis of risk of infection by antibody status (entry date 28 days following first antibody test for all participants): restricted to primary sample (n=2,111)**

|  | **Stratified by LTCF** | | **Stratified by region** | |
| --- | --- | --- | --- | --- |
|  | **aHR [95% CI]** | **p-value** | **aHR [95% CI]** | **p-value** |
| **Residents** | 0·28 [0·12-0·65] | p=0·003 | 0·17 [0·07-0·39] | p<0·001 |
| **Staff** | 0·67 [0·37-1·22] | p=0·193 | 0·45 [0·24-0·83] | p=0·010 |

aHR adjusted for age and gender

**Table S4**: **Multivariate analysis of risk of infection by antibody status (entry date 28 days following first antibody test for all participants): full sample (n=2,220)**

|  | **Stratified by LTCF** | | **Stratified by region** | |
| --- | --- | --- | --- | --- |
|  | **aHR [95% CI]** | **p-value** | **aHR [95% CI]** | **p-value** |
| **Residents** | 0·28 [0·12-0·65] | p=0·003 | 0·17 [0·07-0·39] | p<0·001 |
| **Staff** | 0·64 [0·35-1·16] | p=0·142 | 0·44 [0·24-0·82] | p=0·009 |

aHR adjusted for age and gender

### **STROBE statement**

|  | Item No | Recommendation | Page No |
| --- | --- | --- | --- |
| **Title and abstract** | | |  |
|  | 1 | (*a*) Indicate the study's design with a commonly used term in the title or the abstract | 2 (abstract),  4 (methods) |
|  |  | (*b*) Provide in the abstract an informative and balanced summary of what was done and what was found | 2 |
| **Introduction** | | |  |
| Background/rationale | 2 | Explain the scientific background and rationale for the investigation being reported | 4 (background) |
| Objectives | 3 | State specific objectives, including any prespecified hypotheses | 4 (background) |
| **Methods** | | |  |
| Study design | 4 | Present key elements of study design early in the paper | 4-6 (methods) |
| Setting | 5 | Describe the setting, locations, and relevant dates, including periods of recruitment, exposure, follow-up, and data collection | 4-5 (methods) |
| Participants | 6 | *a) Cohort study*? Give the eligibility criteria, and the sources and methods of selection of participants. Describe methods of follow-up  *Case-control study*? Give the eligibility criteria, and the sources and methods of case ascertainment and control selection. Give the rationale for the choice of cases and controls  *Cross sectional study*? Give the eligibility criteria, and the sources and methods of selection of participants | 4-6 (methods) |
|  |  | (*b*) *Cohort study*? For matched studies, give matching criteria and number of exposed and unexposed  *Case-control study*? For matched studies, give matching criteria and the number of controls per case | 6 (methods) |
| Variables | 7 | Clearly define all outcomes, exposures, predictors, potential confounders, and effect modifiers. Give diagnostic criteria, if applicable | 5-7 (methods) |
| Data sources/ measurement | 8* | For each variable of interest, give sources of data and details of methods of assessment (measurement). Describe comparability of assessment methods if there is more than one group | 5-7 (methods) |
| Bias | 9 | Describe any efforts to address potential sources of bias | 6 (methods) |
| Study size | 10 | Explain how the study size was arrived at | 4-5 (methods) |
| Quantitative variables | 11 | Explain how quantitative variables were handled in the analyses. If applicable, describe which groupings were chosen and why | 5-7 (methods) |
| Statistical methods | 12 | (*a*) Describe all statistical methods, including those used to control for confounding | 5-7 (methods) |
|  |  | (*b*) Describe any methods used to examine subgroups and interactions | 5-7 (methods) |
|  |  | (*c*) Explain how missing data were addressed | 5-7 (methods) |
|  |  | (*d*) *Cohort study*? If applicable, explain how loss to follow-up was addressed  *Case-control study*? If applicable, explain how matching of cases and controls was addressed  *Cross sectional study*? If applicable, describe analytical methods taking account of sampling strategy | 5-7 (methods) |
|  |  | (*e*) Describe any sensitivity analyses | 7 (methods) |
| **Results** | | |  |
| Participants | 13* | (*a*) Report numbers of individuals at each stage of study? eg numbers potentially eligible, examined for eligibility, confirmed eligible, included in the study, completing follow-up, and analysed | 8 (results), supplement |
|  |  | (*b*) Give reasons for non-participation at each stage | 8 (results), supplement |
|  |  | (*c*) Consider use of a flow diagram | supplement |
| Descriptive data | 14* | (*a*)Give characteristics of study participants (eg demographic, clinical, social) and information on exposures and potential confounders | 8, table 1 |
|  |  | (*b*) Indicate number of participants with missing data for each variable of interest | 8, table 1 |
|  |  | (*c*) *Cohort study*? Summarise follow-up time (eg average and total amount) | 8 (results), figure 1 |
| Outcome data | 15* | *Cohort study*? Report numbers of outcome events or summary measures over time | 8 (results), table 2, figure 1 |
|  |  | *Case-control study?* Report numbers in each exposure category, or summary measures of exposure | n/a |
|  |  | *Cross sectional study?* Report numbers of outcome events or summary measures | n/a |
| Main results | 16 | (*a*) Report the numbers of individuals at each stage of the study?eg numbers potentially eligible, examined for eligibility, confirmed eligible, included in the study, completing follow-up, and analysed | 8 (results), supplement |
|  |  | (*b*) Give reasons for non-participation at each stage | 8 (results), supplement |
|  |  | (*c*) Consider use of a flow diagram | supplement |
| Other analyses | 17 | Report other analyses done?eg analyses of subgroups and interactions, and sensitivity analyses | 8-9 (results), tables 3-4, supplement |
| **Discussion** | | |  |
| Key results | 18 | Summarise key results with reference to study objectives | 9-11 (discussion) |
| Limitations | 19 | Discuss limitations of the study, taking into account sources of potential bias or imprecision. Discuss both direction and magnitude of any potential bias | 9-11 (discussion) |
| Interpretation | 20 | Give a cautious overall interpretation of results considering objectives, limitations, multiplicity of analyses, results from similar studies, and other relevant evidence | 9-11 (discussion) |
| Generalisability | 21 | Discuss the generalisability (external validity) of the study results | 9-11 (discussion) |
| **Other information** | | |  |
| Funding | 22 | Give the source of funding and the role of the funders for the present study and, if applicable, for the original study on which the present article is based | 7 (role of funding source) |
